# Group-derived and individual disconnection in stroke: recovery prediction and deep graph learning

**DOI:** 10.1101/2025.07.02.25330753

**Authors:** Patrik Bey, Kiret Dhindsa, Torsten Rackoll, Jan Feldheim, Marlene Bönstrup, Götz Thomalla, Robert Schulz, Bastian Cheng, Christian Gerloff, Matthias Endres, Alexander Heinrich Nave, Petra Ritter

## Abstract

Recent advances in the treatment of acute ischemic stroke contribute to improved patient outcomes, yet the mechanisms driving long-term disease trajectory are not well-understood. Current trends in the literature emphasize the distributed disruptive impact of stroke lesions on brain network organization. While most studies use population-derived data to investigate lesion interference on healthy tissue, the potential for individualized treatment strategies remains underexplored due to a lack of availability and effective utilization of the necessary clinical imaging data. To validate the potential for individualized patient evaluation, we explored and compared the differential information in network models based on normative and individual data. We further present our novel deep learning approach providing usable and accurate estimates of individual stroke impact utilizing minimal imaging data, thus bridging the data gap hindering individualized treatment planning.

We created normative and individual disconnectomes for each of 78 patients (mean age 65.1 years, 32 females) from two independent cohort studies. MRI data and Barthel Index, as a measure of activities of daily living, were collected in the acute and early sub-acute phase after stroke (baseline) and at three months post stroke incident. Disconnectomes were subsequently described using 12 network metrics, including clustering coefficient and transitivity. Metrics were first compared between disconnectomes and further utilized as features in a classifier to predict a patients’ disease trajectory, as defined by three months Barthel Index. We then developed a deep learning architecture based on graph convolution and trained it to predict properties of the individual disconnectomes from the normative disconnectomes.

Both disconnectomes showed statistically significant differences in topology and predictive power. Normative disconnectomes included a statistically significant larger number of connections (N=604 for normative *versus* N=210 for individual) and agreement between network properties ranged from *r*^2^=0.01 for *clustering coefficient* to *r*^2^=0.8 for *assortativity,* highlighting the impact of disconnectome choice on subsequent analysis. To predict patient deficit severity, individual data achieved an AUC score of 0.94 compared to an AUC score of 0.85 for normative based features. Our deep learning estimates showed high correlation with individual features (mean *r*^2^=0.94) and a comparable performance with an AUC score of 0.93.

We were able to show how normative data-based analysis of stroke disconnections provides limited information regarding patient recovery. In contrast, individual data provided higher prognostic precision. We presented a novel approach to curb the need for individual data while retaining most of the differential information encoding individual patient disease trajectory.

## Introduction

The detrimental effect of ischemic stroke on patient quality-of-life is well established.^1^ Activities of daily living (ADL) for example are often impacted by a range of post-stroke disabilities, such as impaired motor function, with a high prevalence of long term implications for both patients and care-givers.^2^ Extensive studies have investigated contributing factors, from lesion topology and location to alterations in both functional and structural connectivity^3–6^, with the goal of identifying feasible treatment targets. However, reliable biomarkers for predicting individual disease trajectory after stroke to guide treatment regimen remain an ongoing challenge, particularly during the acute phase following stroke.^7–9^ Network effects of stroke in particular have become an active focus of investigation to understand the distributed impact of focal lesions.^10,11^ These efforts introduced novel methodologies like the lesion network mapping approach^12,13^ which was subsequently further developed to introduce the disconnectome.^14^ Current literature focuses on one of two major characteristics of lesion impact. The first group of studies investigates the network impact of stroke via an estimated model of the lesion-compromised white matter tracts. The second group investigates functional network disruptions of stroke using functional magnetic resonance imaging (MRI) data.^5,15,16^ Both approaches frequently follow a population-based, or normative, modelling approach for the underlying data. Such disconnectomes have been shown to relate to brain function in health and disease^14^ and have enabled prediction of various clinical and neuropsychological scores with moderate to good accuracy.^6,9,17–19^

They have, however, fallen short of providing a feasible clinical application. At the same time, only a small number of studies have focused on the estimation of individual structural disconnectomes.^20^ This fact is due to two major aspects that drive the development of normative connectomics. The first aspect represents an understanding that the study of disruption of normal healthy signal represents a feasible approximation to the disease induced alterations of the individual. The other main driver is the amplified requirements on data acquisition as well as subsequent data processing necessary to facilitate individual connectomics. The clinical context of stroke patient MRI data introduces strong confines to the duration of scanning protocols, due to the importance of fast differential diagnosis and avoidance of additional burdens on the patient. Henceforth, the acquisition of high-quality MRI acquisition sequences is not feasible with the current clinical neuroimaging infrastructure. Previous work, however, has demonstrated limitations in the normative approach for the investigation of individual disease trajectories caused by brain pathologies.^21^ These findings suggested differences in the encoded information relating to individual patient outcome between networks estimated from normative or population data and network analysis utilizing individually acquired patient MRI data, suggesting a trade-off between approaches is required to allow for prospective clinical utility. To carefully investigate these differences and provide a comprehensive comparison, in this study, we evaluated the relationship between the normative and individual approach by creating both a normative as well as an individual disconnectome for each stroke patient. We hypothesized, the differences between both approaches to be driven by methodological limitations inherent in the creation of population templates, in particular strong agreements on subcortical brain areas between participants with a decrease in individual sensitivity towards cortical areas, as shown for the creation of anatomical templates.^22^ This loss of individuality carries over in the establishment of population tractograms, as utilized when creating normative disconnectomes. This leads to an increased presence of robust long distance white matter tracts, compared to shorter and weaker individually driven connectivity between cortical regions-of-interest (ROI), potentially reducing subject specificity in localized connection-based analysis, as in the case of lesion disconnectomes. Henceforth, refining our understanding of such differences provides major improvements in modelling of long-term patient outcomes and enables potential clinical applications of such disease outcome models. To this end we utilized DWI MRI data of a total of *N*=78 acute to sub-acute stroke patients from two data cohorts. The severity of stroke based disabilities was assessed via Barthel Index (BI)^23^, a single measure capturing comprehensive estimates of ADL and a patients capability to perform everyday tasks and whose improvement over time directly translates into an increase in patient quality of life.^24^ Disease trajectory was subsequently evaluated using the three-months post stroke BI for each patient. In the first section of this manuscript, we describe the resulting disconnectomes using various network measures capturing a wide range of properties relating to the functional integration and spatial propagation of the lesion impact network. We compare the disconnectome pairs for each patient to explore occurring differences and highlight the varying descriptions of both approaches. In a subsequent validation step we used the network measures as input features to a non-linear support vector machine (SVM) machine learning model to probe the encoded differential information of both approaches in relation to the patient’s long-term disease trajectory. In the second part of this study, we built upon the prior analysis of both disconnectome approaches and introduced a novel individual disease trajectory modelling approach as a proof-of-concept for clinical applications. Hence we employed the recently developed novel deep learning (DL) tool of graph convolutional neural networks (GCN).^25^ Following previous work on functional MRI^26^, here we implemented such an architecture for use with structural networks, in particular disconnectomes. This advancement allowed for the estimation of individual network properties from the normative input disconnectome of a patient, therefore integrating a hybrid model of both approaches without the need for extensive imaging modalities as required for the individual approach. The resulting estimates of network properties were afterwards once more used as input features for the same SVM classification task as in the previous validation step. The full analysis workflow is displayed in Fig. 1. To accurately portray patient disease severity and corresponding three-month trajectory levels and validate the current model, we defined classes of patient deficit severity from the corresponding BI scores following Lee et al.^27^. The introduced groups were concatenated to represent three classes of distinct severity levels: *severe*, *moderate*, and *mild*. Fig. 2A shows recovery groups with corresponding BI scores for all included patients from baseline to three-months post stroke insult. The current work provides the first extensive investigation of differences between normative and individual network effects of stroke and their relationship to long-term patient outcome. Most importantly our work further introduces a novel DL framework, building upon data routinely available in the clinical context of stroke, that provides performance in individual patient disease trajectory prediction feasible for integration into clinical routine.

**Figure 1.**
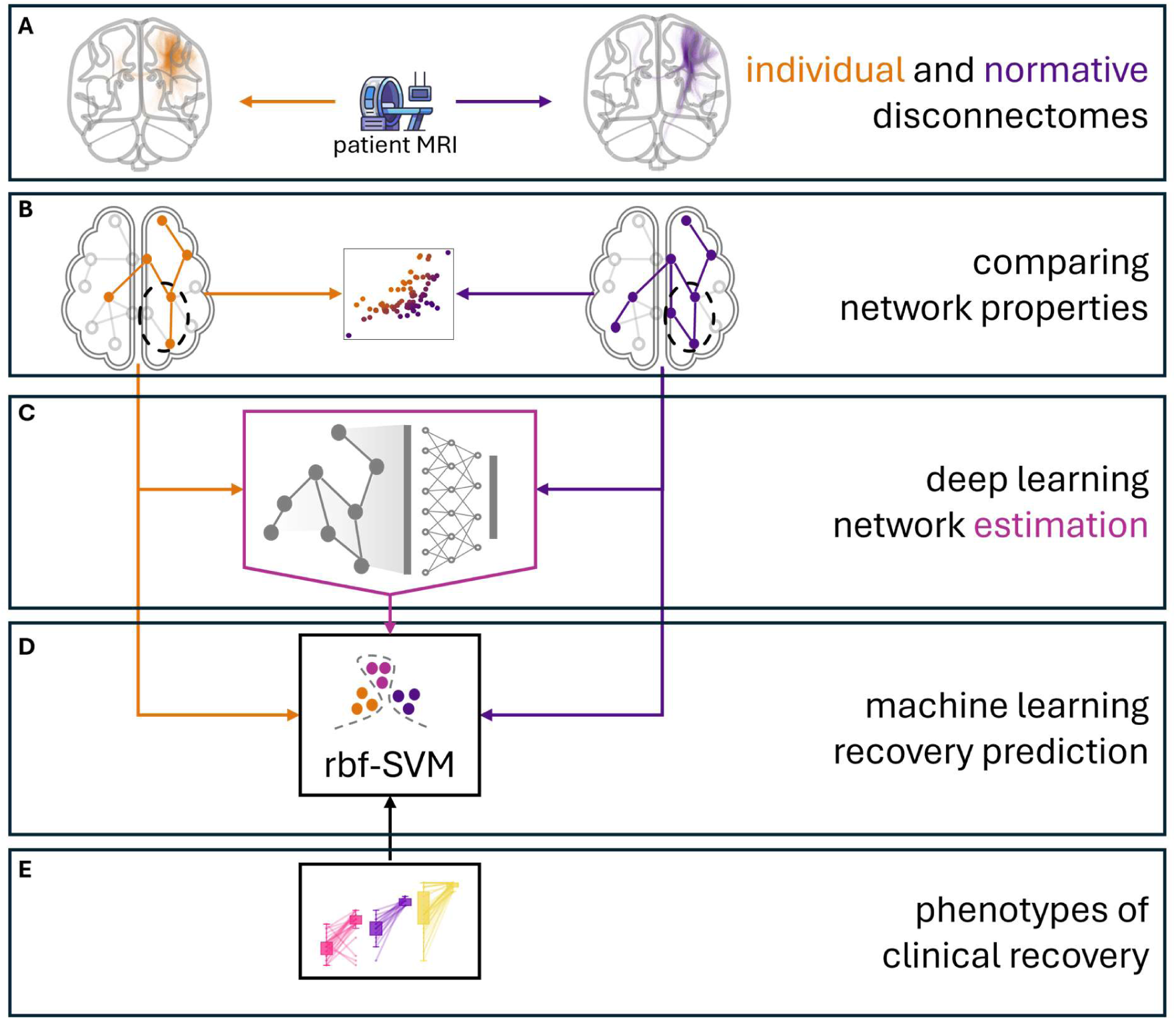
Analysis workflow of the present study. Based on patient MRI data, two sets of disconnectomes were created (**A**). The normative (right) disconnectome and the individual disconnectome (left). Both sets of graphs (**B**) were subsequently described via a set of twelve network metrics. In addition, a graph convolutional neural network was trained taking the normative disconnectome graph as input and learning a mapping to the individual network metrics (**C**). All three resulting sets of network measures were used as input features to SVM classifiers (**D**) with a radial basis function (rbf) non-linear kernel function, to predict patient labels of disease trajectory from baseline to three-months post stroke incident (**E**).

**Figure 2.**
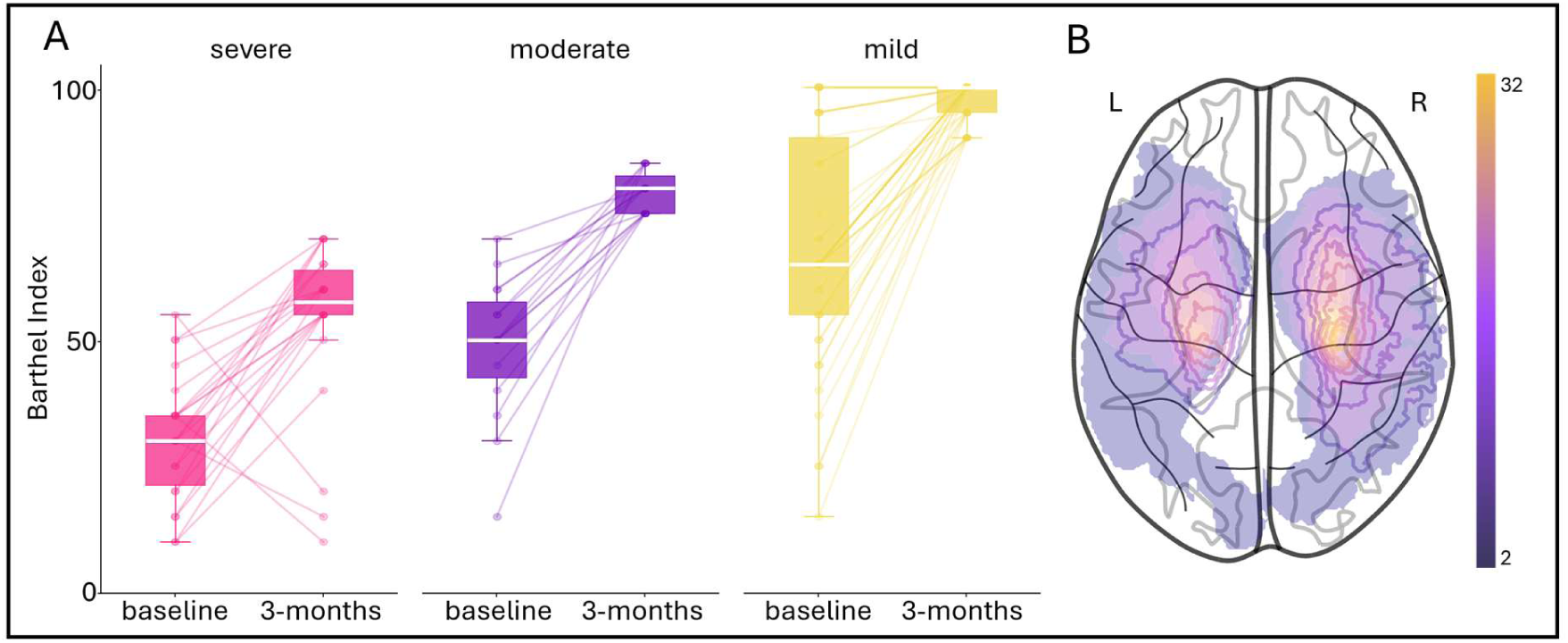
Clinical phenotypes of disability for all patients. (**A**) Patients BI scores at both baseline and three-months post stroke. The patients are divided into corresponding severity groups at three-months long term stages. The scatter plots at baseline are color-coded according to the initial severity class detailing the individual disease trajectory from baseline to late sub-acute stages. The right panel (**B**) shows the average lesion mask of the patient population utilized in this study after registration of all binary lesion masks to MNI152 standard space.

## Materials and methods

### Datasets

The data used in this study consist of a total of *N*=78 ischemic stroke patients from two different cohorts summarized in Table 1. The first data set was acquired at Universitätsklinikum Hamburg-Eppendorf (UKE), Hamburg, Germany after approval of the corresponding ethics board (PV3777) and has previously contributed to a wide range of studies.^15,28,29^ The second data set is comprised of patients of the BAPTISe^30^ study, a subset of the PHYS-Stroke^31^ randomised-controlled trial, acquired at Charité Universitätsmedizin Berlin, in Berlin, Germany and approved by the corresponding ethics board (EA1/137/13). The present patient data were selected from the corresponding larger cohorts in fulfilment of the processing and modelling requirements, as defined in Supplementary Table 1. These data requirements led to a significant reduction in patient inclusion compared to the original size of both data cohorts. The present study was further approved by the ethics committee of Charité Universitätsmedizin Berlin (EA1/309/24). Included MRI modalities were structural T1w and FLAIR images as well as DWI based tractography.

**Table 1.** Dataset descriptor for both included data cohorts. Brackets contain the standard deviation where applicable.

| Data set | UKE | BAPTISe |
| --- | --- | --- |
| Sample size | 24 | 54 |
| age | 60.5 | 67.1 |
| sex | 10 females | 22 females |
| Stroke time (baseline) | 4 ($\pm 1$) days post stroke | 24 ($\pm 11$) days post stroke* |
| Stroke time (sub-acute) | 90 ($\pm 5$) days post stroke | 91 ($\pm 11$) days post stroke |
| Median BI (baseline) | 85 ($\pm 27$) | 50 ($\pm 18$) |
| Median BI (sub-acute) | 100 ($\pm 13$) | 80 ($\pm 22$) |
| Median NIHSS (baseline) | 4.5 ($\pm 3.3$) | 10 ($\pm 4.7$) |
| Median MRS (baseline) | 3 ($\pm 1.4$) | 3 ($\pm 0.95$) |
| Lesion volume (baseline) | 5.655mL ($\pm 31$mL) | 8.89mL ($\pm 45$mL) |
| Lesion hemisphere | 50% left | 41% left |

BI = Barthel Index, NIHSS = National Institute of Health Stroke Scale, MRS = Modified Rankin Scale

#### Phenotyping

The clinical expression of stroke induced disabilities was assessed using a range of neuropsychological scores, commonly employed in clinical diagnosis. To enable harmonization across the included data sets, the present analysis focuses on the BI as a well-established score for the assessment of ADL.^23,27^ As the current study aimed to predict stroke trajectory for patients individually, the BI scores were divided into three classes representing *minor*, *mild*, and *severe* disabilities extending upon previously introduced groupings.^27,32^ Patients were henceforth assigned *severe* class membership with a BI equal to or below 70, *moderate* class membership with a BI below 90, and *mild* class membership otherwise. These labels allowed for multi-class classification of the three months disease trajectory for each patient. The underlying stroke lesions at the baseline stage are shown in Fig. 2B displaying a common pattern of ischemic stroke patient populations.^14^

### Processing

#### MRI processing

The multimodal MRI data of this study was processed using a range of established and automated processing frameworks including LeAPP^29^ and FastSurfer^33^ specifically adjusted for stroke patients, to minimize distortions potentially influencing downstream analysis^34,35^ and is further detailed in supplementary material processing framework. Disconnectome creation was done twofold as described below. Both disconnectomes are constructed via the combination of a subset of tracts from the varying tractograms and a brain parcellation mask. The final disconnectome network is then defined via an adjacency matrix with zeros for non-existing connections within the lesion network and a positive value representing the count of tracts between brain ROI pairs.

#### Normative disconnectome creation

Following current literature on lesion-based connectivity networks^18,19^, we estimated the normative disconnectomes using a population-derived tractogram, in this case a normative tractogram created based on 985 Human Connectome Project (HCP)^36^ healthy participants^37^ (median age 31.6 years). To this end we projected the patient lesion masks in MNI152 space, as performed during the processing of the individual multimodal MRI data, into the tractogram and extracted all tracts intersecting the binary lesion volume using *MRTrix3*^38^ as implemented in LeAPP and previously discussed.^39,40^

#### Individual disconnectome creation

Creating the individual disconnectome was again based on the LeAPP processing framework and followed the same approach as the normative disconnectome. However, in this instance the native patient lesion mask was embedded in the individual patient tractogram, also containing a total of 10 million estimated tracts matching the dimensionality of the normative tractogram, creating a lesion network based solely on individual patient data.

#### Network metrics

To capture a broad range of network properties that might encode complex clinical information regarding the underlying mechanisms of stroke recovery, we computed the following twelve network measures *node strength, clustering coefficient, centrality, assortativity, density, shortest path length, non-randomness, transitivity, diameter, periphery size, bridge count* and *independents count.*^41,42^ These network measures capture various properties relating to the functional integration and spatial propagation of the underlying lesion impact network. The measure *density* for example encodes information about the relationship between ROIs as it relates to the existing number of pathways relative to the theoretically possible number of pathways providing information about the cost of information processing within a network. Such information may allow for inference on the functional integration of the lesion network within the overall information processing network of the brain.^41^ Additionally, measures such as *periphery* encode information about the spatial and hierarchical distribution of nodes relative to one another, as it captures the number of ROIs on the outskirt of the network.^43^ A measure that transfers information on the localization of the spatial impact of the lesion on the underlying pathways. Computation of these measures was performed after thresholding and binarization of the disconnectome graphs. To reduce the chance of false positive connections and retain robust tract estimates only the top ten percent of edges in connectivity strength were kept in the graph. Network measures were computed for both disconnectomes for each patient. A more detailed description of all network measures can be found in Supplementary Table 2.

#### Graph comparison

To further investigate differences between individual und normative disconnectomes we computed the spectral distance^44^ between each disconnectome pair. The spectral distance measure captures structural differences between graphs, in this case the disconnectomes. A secondary use of the spectral distance measure was to allow for the investigation into the impact of lesion volume on the differences between both disconnectome approaches.

### Machine learning

To investigate this study’s hypothesis on the difference in encoded information in normative versus individual disconnectomes and the potential to create individual information from the normative data, we deployed two machine learning based frameworks.

The first application utilized SVM for the classification of stroke recovery degree for each patient, based on disconnectome network properties. The second framework is a deep GCN model, attuned to learn a representation mapping from the normative disconnectome of a patient to its individual disconnectome characteristics. Both methodologies were employed using a leave-one-out cross validation (LOOCV) scheme to ensure generalizability and avoid overfitting while maximizing the available training data in each instance. The implementations were done using the Python programming language, in particular libraries such as *numpy*, *scikit-learn*, *PyTorch* and *PyTorch-Geometric*. Computations were performed on a Ryzen 9 5900x CPU with 64 GB of RAM and a Nvidia RTX 4070 super GPU with 12GB of memory.

#### Stroke recovery classification

The prediction of disability class membership for a given patient was performed using a non-linear SVM. SVM is a maximum margin classifier that identifies a separating hyperplane between data samples in a higher dimensional representation of the input feature space.^45^ The distance of a subset of data samples, the so-called support vectors, to the hyperplane drives the profile of the plane, weighted by the adjustable *cost* parameter. The mapping of the input data to the high dimensional space is done utilizing a radial basis function (rbf) kernel with a corresponding *gamma* parameter, defining the shape of the space representation.

Hyperparameter tuning was performed using a grid search approach for both parameters with values ranging from 0.0001 to 100, resulting in a total of 441 combinations, each undergoing a full LOOCV run. The input features were whitened before classification to harmonize the varying scales present in the network features and to remove potential bias sources for the classifier. The overall performance of a given model was evaluated using a weighted F1 score to incorporate the different class sizes in the performance metric as well as receiver operating characteristics area under the curve (AUC) highlighting potential disparities in class predictions.

#### Feature importance

After identification of the optimal parameter combination for the given model, permutation feature importance (PFI) was computed to assess each feature’s contribution to the overall classification accuracy. To this end the model was trained using the optimal hyperparameters, while a single feature was randomly permuted (i.e., it’s values were shuffled). The test set was predicted and the resulting F1 score computed. A total of 50 permutations were performed for each feature during each LOOCV run. The mean F1 score was computed for each permuted feature and the difference to the previously achieved F1 scores, without permutation, was calculated. This approach created an average importance for each feature defined via the overall change in classification performance. For further investigation of the PFI values, z-scores were computed for each given feature set allowing for comparison across classification models.

#### Deep estimation of individual information

We further developed and implemented a GCN model consisting of both graph convolution and linear layers for the task of estimating individual network properties from normative graph input. In the first module the graph input information is projected into higher dimensional graph embeddings for each node while continuously reducing graph size, e.g. node count. The initial node embeddings for each graph are given via the node degree of each node of the input graph, as a basic measure of ROI integration within the given lesion network without increasing computational cost of the initialization step. The last step of the first module creates a 256-dimensional graph embedding vector, which in turn serves as input to the second module. The linear layers of the second module reduce the dimensionality to the final output dimension using linear transformations of the input tensor with the learned weight matrices, matching the 12-dimensional vector of the target values representing the 12 individual network metrics. The *SAGEConv*^46^ convolutional layers were implemented in pairs with a matching T*opKPooling*^47,48^ layer as shown in Fig. 3. A total of four such layer blocks were implemented with resulting graph embedding dimensions of 32, 64, 128 and 256. While the initial node embeddings for each node are given by its degree, the succeeding graph embeddings are updated recursively within each GCN layer using a *message passing* range of six, leading to the integration of information from neighbouring nodes with a maximum distance of six into the embedding for each given node. An overall dropout rate of 0.1 was employed resulting in the removal of random 10% of connections during graph convolution and ensure stability of model conversion and generalizability. *TopKPooling* then reduces the overall size of the graph by dropping nodes based on a softmax evaluation of the learned projection score with layer wise decreasing drop rates of 0.8, 0.6, 0.4, and 0.2 for the final layer. In the second module the linear layers reduce the graph embedding vector recursively from 256 via output dimensions of 128,64,32 to the final 12-dimensional estimates. For both modules activation at the respective layers were computed using the Leaky Rectified Linear Unit (ReLU) activation function. The final model output vector is evaluated using the mean square error (MSE) loss function. Subsequently the used Adam optimizer with a *learning rate* of 0.01 and a *weight decay* factor of 1e^−4^ is updated to complete a single epoch. A total of 50 such epochs are run within each LOOCV loop. Training of the network was performed using a batch size of *N*-1 incorporating the full training data set within each batch due to low memory requirements given by the graph representation of the input data. Following Goodhart’s law, for validation purposes we used *R2* scores to assess testing performance to avoid circular analysis by utilizing the same measure during testing as during training. The final count of learnable weights for the introduced GCN model was 1091180, representing the moderately low complexity of the model compared to current deep learning architectures, to accommodate for the relatively low sample size and minimize the risk of overfitting.

**Figure 3.**
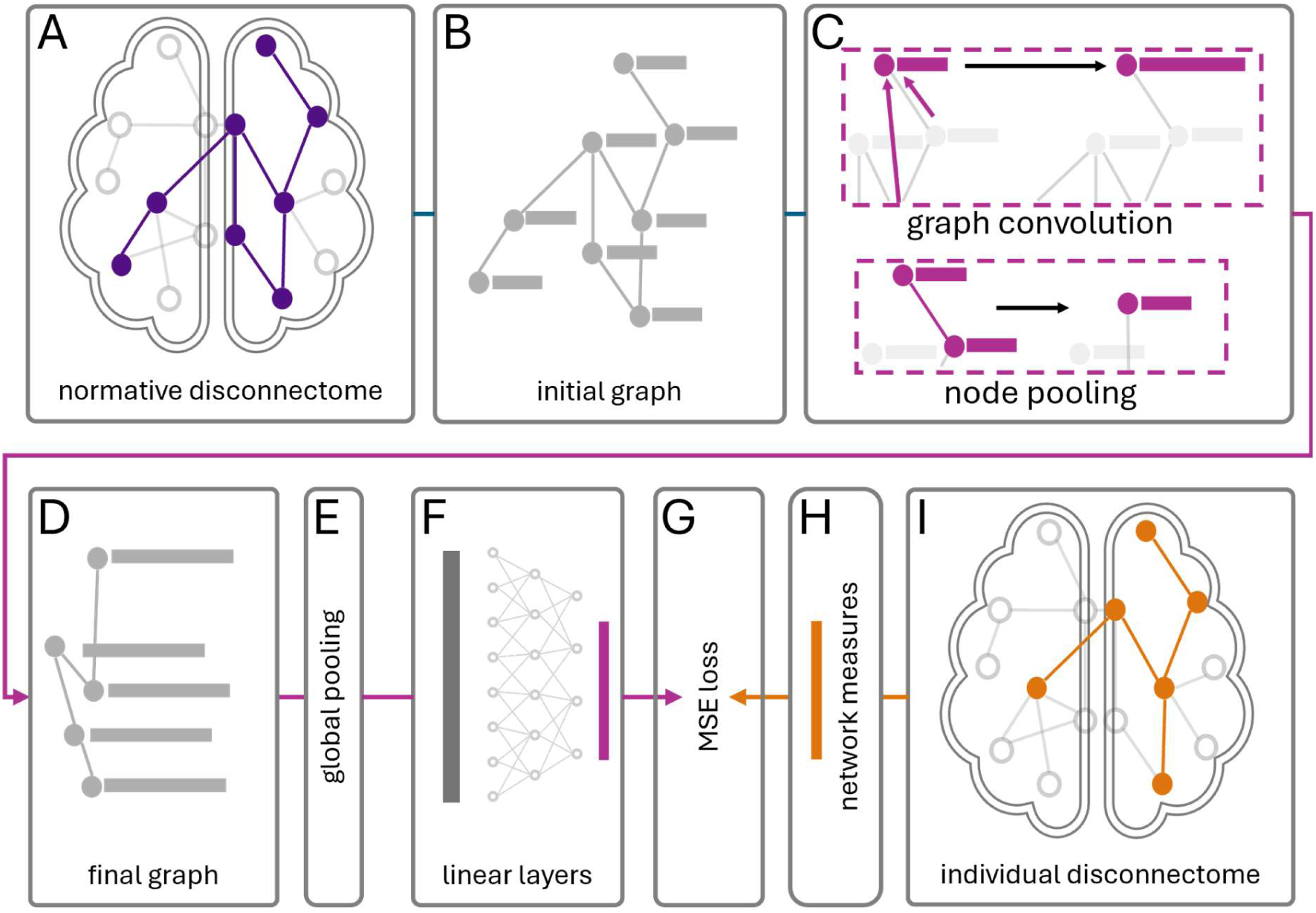
Graph Convolution Model. architecture as implemented in this study. The normative input graphs (**A**) are initialized with node degree as corresponding node feature and stored as graph representations (**B**). Each graph is then forwarded through a total of four GCN layer pairs (**C**), made up of first a GraphSAGE convolution layer utilizing message passing step size of 6, integrating neighbouring and adjoining nodes. The final graph embeddings are then pooled using SoftMax and a decreasing pooling ratio. This process is repeated through all four GCN layers. The final graph representation (**D**) is then pooled via global max pooling to create a single 256-dimensional graph representation (**E**). This representation is passed through four fully connected linear layers with ReLU activation, reducing the dimensionality to 12. The output is evaluated against the true individual disconnectome (**I**) network measures (**H**) using a mean squared error (MSE) loss function (**G**).

## Results

### Normative and individual disconnectome dissimilarities

The networks created using first the population-based normative tractograms and second the individual patient DWI data, show statistically significant differences in both whole brain integration and local properties. Fig. 4A shows an average disconnectome from either approach for the same group of subjects with an overlapping lesion location in left hemisphere insula and frontal opercular area. Normative tractogram based disconnectomes (left) show a stronger integration of contralesional and homotopic connections, which have been shown to decode information of disease severity and recovery in functional MRI.^49–51^ While individual disconnectomes show a significantly reduced overall connectivity (on average reduced by a factor of 2.9, see Table 2). Differences between both approaches were significant across multiple properties such as the overall count of ROI in the present network, the count of both ipsilesional and contralesional ROIs as well as the overall count of subcortical ROIs integrated as part of the lesion disrupted connectivity of a given patient. These findings align with the general interpretation of normative population data to focus on population level agreement of stronger connections, while individual data might retain a more fine-grained representation of potentially weaker connection, that are potentially lost in population-based data due to the reduced specificity of cortical areas in group averages^22^.

**Figure 4.**
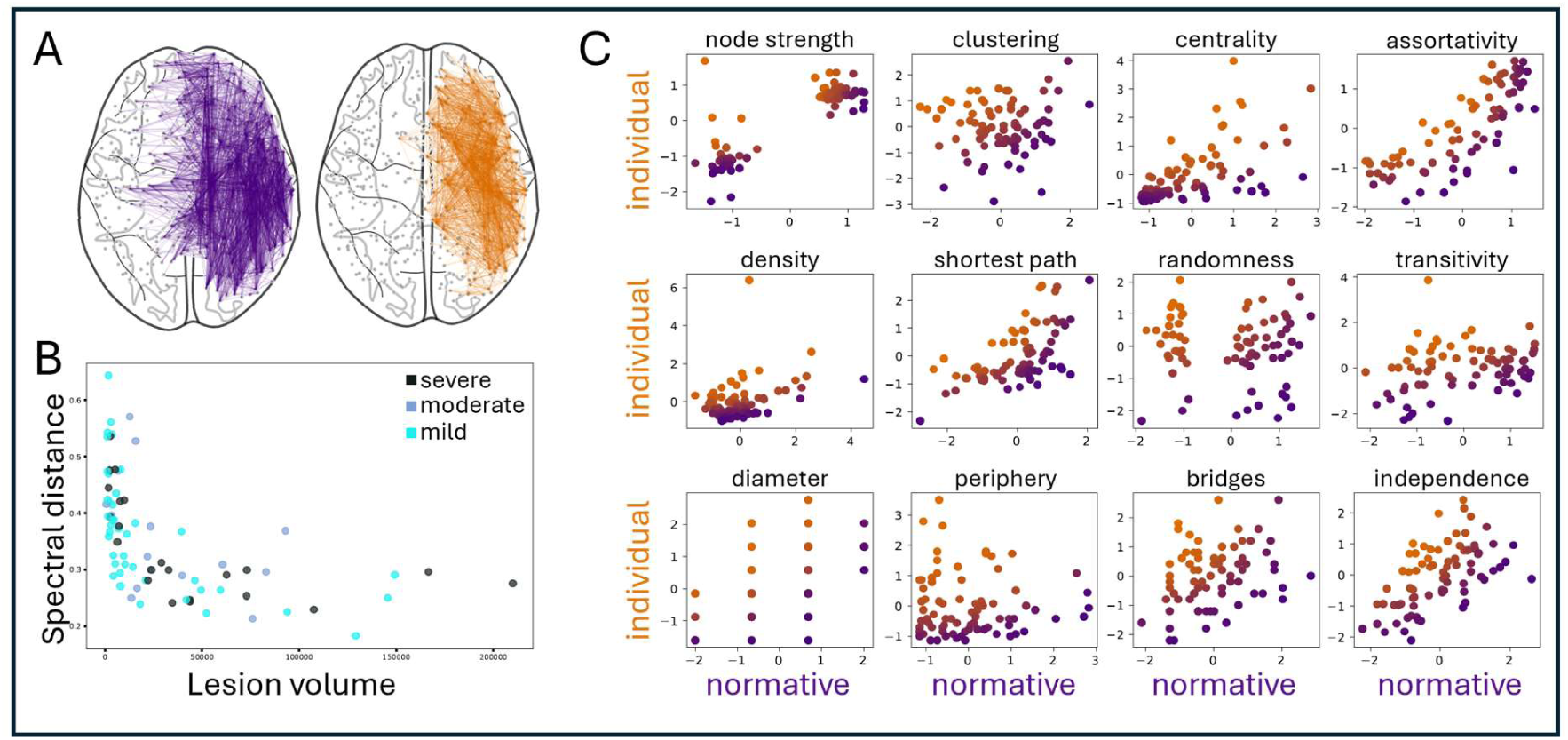
Disconnectome comparison between the normative and individual approach. (**A**) Displays the average disconnectome for a subset of patients with similar lesion location (*N*=14 patients with left hemisphere insula and frontal opercular area lesion). The left graph showing the normative disconnectome while the right shows the average individual disconnectome. The spectral distance capturing structural differences in the underlying graphs between both approaches is shown in (**B**). The spectral distance is shown in relationship to the corresponding patient’s lesion size, highlighting a clear negative relationship between both properties. (**C**) Shows the differences between all network properties of both approaches. Half of all network properties show a significant correlation between both approaches, including node strength, centrality, assortativity, shortest path length, diameter, and average independents count. The other half of features highlight the varying degrees of information encoded via the disconnectome approach, including clustering, density, non-randomness, transitivity, periphery size, as well as bridge count.

**Table 2.** Disconnectome comparison for basis statistics. Values represent median values (± standard deviation) for both disconnectomes. Significant differences were evaluated using a related two-sided T-test with an *a*=0.05 significance cutoff.

| Approach | Connection count | Total ROI count | Ipsilesion ROI count | Contralesion ROI count | Subcortical ROI count |
| --- | --- | --- | --- | --- | --- |
| Normative | 604 (± 443) | 65 (± 47) | 40 (± 38.1) | 12 (± 9.1) | 72 (± 36.6) |
| Individual | 210 (± 181) | 36 (± 28) | 26 (± 26.7) | 0 (± 3.0) | 41 (± 19.3) |
| p-value | $1.3e^{-19*}$ | $2.4e^{-15*}$ | $1.8e^{-8*}$ | $8.3e^{-18*}$ | $5.1e^{-22*}$ |
| t-statistic | 12.2 | 2.9 | 6.3 | 11.2 | 13.5 |

ROI = Region of interest

The overall difference in graph structure between a patients’ disconnectome pair was assessed using spectral distance. Fig. 4B shows the negative non-linear relationship between lesion volume and corresponding spectral distance for each patient. The distance between both disconnectome approaches is therefore only relatively large for smaller lesion volumes while after a certain point lesion volume appears to no longer contribute to differences in disconnectomes. There is furthermore no clear relationship between spectral distance or lesion volume to disease trajectory as validated using ANOVA (spectral distance *p-value*=0.68 and *statistic*=0.4, lesion volume *p-value*=0.22 and *statistic*=1.5) and highlighted by the lack of separation along class lines in the colour coding of the plot, highlighting the complex nature of information encoding disease trajectory beyond lesion volume. The relationship between both disconnectome approaches with regards to individual network parameters is shown in Fig. 4C. While half of all features show a consistent relationship across disconnectomes (node strength *r*^2^=0.68, centrality r2=0.65, assortativity *r*^2^=0.8, shortest path *r*^2^=0.61, diameter *r*^2^=0.55, and independence *r*^2^=0.59), the other half of features (clustering coefficient *r*^2^=-0.01, density *r*^2^ = 0.23, randomness *r*^2^ =0.01, transitivity *r*^2^ =0.25, periphery size *r*^2^=0.02, and bridges *r*^2^=0.28) do not show substantial correlation between disconnectomes.

### GCN disconnectome feature estimates and comparison

The model architecture we introduced in this study was able to create estimates of the individual network metrics while only requiring the standard space aligned lesion mask of a given patient as MRI based input to create the normative disconnectome input, removing the need for high quality DWI data to enable accurate inference of the patients disconnectome. The model achieved a statistically significant reduction in and convergence of the training loss after around 10 epochs (Fig. 5A). The overall performance of the model reached an average R2 score of 0.3 for the test validation and an average MSE score of below 0.05 for the training validation. At the same time the GCN based estimates show strong correlation of all network estimates with individual measures (mean *r*^2^=0.94) is consistent. The reason for the negative correlation however is not clear but consistent across measures. The sign of the correlation is not indicative of the contained differential information regarding the long-term recovery prediction as shown below.

**Figure 5.**
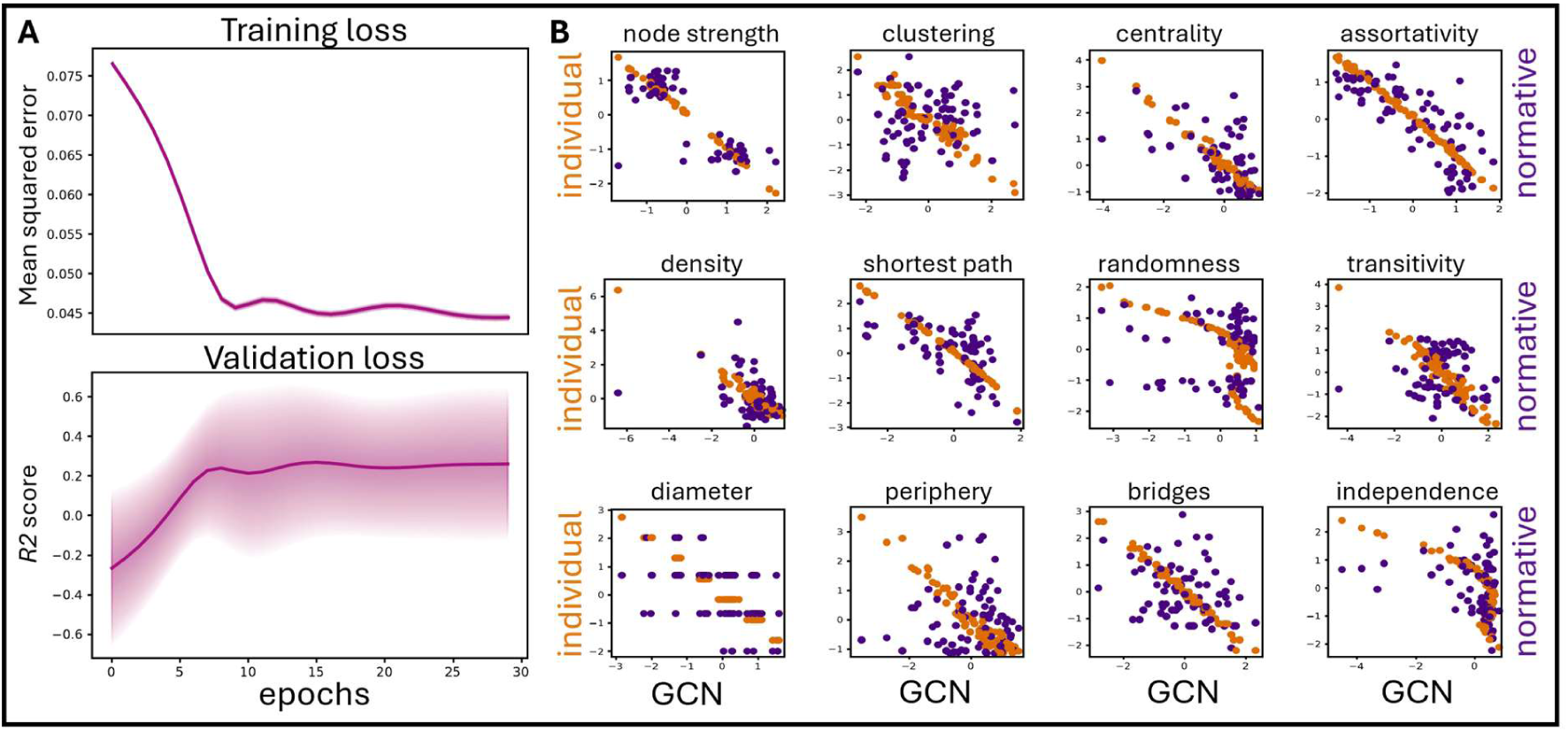
GCN results for both training performance and output network estimates. (**A**) Shows a clear convergence of the model training with a significant decrease in training loss across epochs (top row). The corresponding test performance, as captured by *R2*-score, is shown in the bottom row exhibiting a clear increase in performance as well as a strong variance across LOOCV validation runs with single patients displaying very low R2 scores across all epochs, driving the large variance of the validation loss, as shown via the ribbon around the average value in both plots. (**B**) Shows the corresponding relationship of the predicted network estimates from the GCN model with the computed network features of both disconnectomes. The X-axis represents the corresponding values of the GCN estimates while the Y-axis represents values for both normative and individual disconnectome metrics. As the model was trained to predict individual measures our results show a very strong correlation to these measures with few exceptions, including *independence*, *non-randomness*, and *density*, where a clear hybrid of both models is created.

### Recovery prediction

#### Patient trajectory classification

Our model shows high performance values for the prediction of the three classes of long-term post-stroke disability levels of a given patient. Network features describing properties of the empirically derived individual disconnectome resulted in the highest performance with a weighted *F1* score of 0.95 (*AUC* = 0.94) predicting three-month severity levels. In contrast, normative disconnectome based measures resulted in an *F1* score of 0.89 (*AUC* = 0.85). Our GCN based estimates were further able to approximately reproduce individual disconnectome performance with a *F1* score of 0.94 (*AUC* = 0.93). The most important aspect of this increase in performance from normative to GCN based features lies in the accuracy of predicting the *moderate* recovery group. While all models successfully differentiate patients of the *severe* and *mild* severity classes, the normative features model fails to distinguish *moderate* from *mild* patients by misclassifying 11 out of 15 patients as *mild*. The individual features on the other hand only misclassify 4 out of 15. Moreover, the GCN based estimates accurately predict *moderate* severity for 10 out of the 15 patients in that class reducing the misclassification to 5 from the normative 11. The overall class specific accuracy is shown in Fig. 6A.

**Figure 6.**
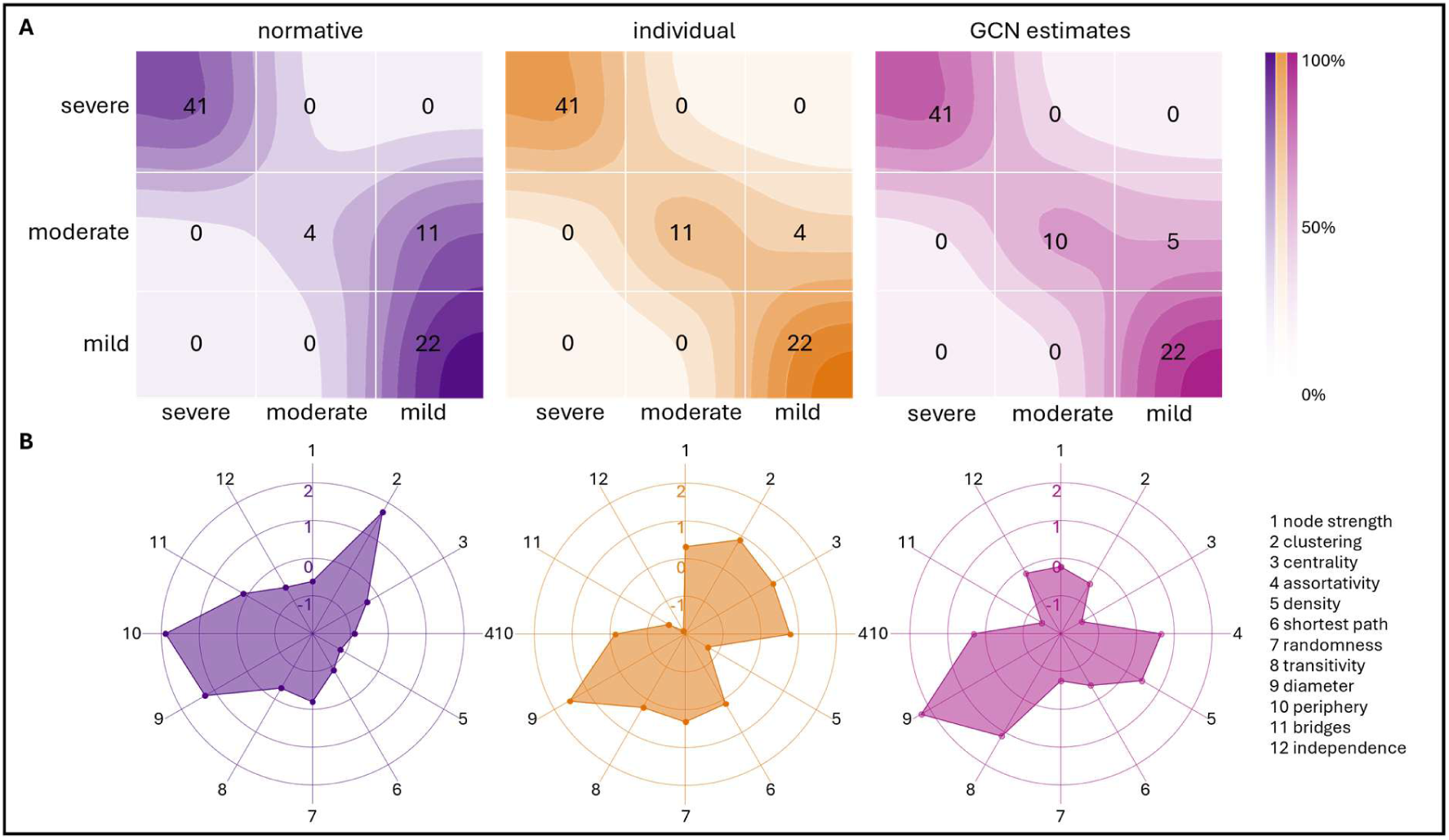
classification performance. (**A**) The differences in performance are highlighted in the confusion matrices for all three optimal classification models. The best performing individual disconnectome features (middle) show the highest accuracy in the classification of the moderate severity class, leading to the high weighted F1 score of 0.95 (AUC = 0.94). The normative model (left) shows a clear limitation to distinguish *moderate* recovery group from the *mild* recovery patients, leading to the lower *F1* score of 0.88 (AUC=0.85). The GCN based model (right) on the other hand almost reaches the same performance exhibited by the individual model by correctly classifying 10/15 moderate recovery patients, leading to an F1 score of 0.94 (AUC = 0.93). The permuted feature importance (PFI) across models for all 12 included network features (**B**). While we see a clear disagreement between normative and individual model there is a strong agreement on feature importance between individual and GCN based model. An interesting find is the importance of hybrid measures (*density*, *randomness*, and *independence*), as given by their relationship to both disconnectomes (see Figure 5), in the case of the GCN model compared to the individual model, showing a strong influence of the hybrid nature to the differential information encoded in the estimates.

#### Disconnectome feature importance

The set of features contributing the most to overall performance changes across underlying feature set. There is a strong disagreement between normative and individual models on the overall importance of features. While there is agreement on the importance of *clustering coefficient*, a measure capturing the overall efficiency in local information transport, even without a strong correlation in this measure between disconnectomes (Fig. 4C), there is a strong disagreement on the importance of *periphery size*. The latter measure captures the overall count of ROIs considered on the outskirt of the created network graph, a property significantly driven by the stronger integration of contralesional and homotopic connections evident in normative disconnectomes compared to individual (see Table 2). Feature importance for the GCN model however resulted in a similar distribution of PFI compared to the individual model with a small number of important variations. The three features that show relatively low correlation from the GCN to the individual model, theorized to present a hybrid estimate between both disconnectomes, led to the strongest disagreements in PFI between the models. Both *density* as well as *independence*, two measures capturing complementary information regarding the tightness of connections in a given graph, show a higher importance for the GCN model. *Randomness* on the other hand is accredited less importance.

## Discussion

Combining both individual and population template MRI data we were able to show, for the first time, how the varying information encoded by either approach, when creating disconnection networks for stroke lesions, drive the expression of biomarkers for stroke patient disease trajectories. Differences were shown to align with the hypothesis that normative data will provide a smoother baseline of connections capturing strong connectivity patterns across major brain connections, while individual data retains a more fine-grained overview even of weaker connections. Building on those results we furthermore introduced a novel hybrid modelling approach using state-of-the-art deep learning methodology to overcome the extensive data requirements of the individual disconnectome approach. Our model was able to achieve comparable performance to the model trained on individual patient data while utilizing only normative input data. This serves as a proof-of-concept for the potential clinical applications of deep learning in supporting stroke patient treatment and recovery decision making.

### Disconnectomes as stroke biomarkers

The information encoded in the estimated disruption of healthy brain connections has previously provided major insights into lesion impact and clinical severity based on disconnection pattern using both normative white matter connections^7,18,52–56^ as well as functional brain networks.^57,58^ At the same time, complimentary works investigating the impact of various normative data sets highlighted the potential for biased analyses using normative data for individual disease-specific inference.^21,59^ Furthermore, additional work has shown how a patient’s individual deviation from the normative control group might drive individual brain topology or disease trajectory^60,61^ in particular in the case of pathological patient brains.^62^ These findings provided a crucial framework for investigating the feasibility of normative disconnectomes in stroke patient recovery and left the neuroimaging community without a scientific consensus on the required data for research into stroke recovery biomarkers. In this study we provided a vital contribution to clarify the resulting differences via an extensive investigation of differences across several scales. The reported differences in baseline statistics of both disconnectomes as shown in Table 2 provide an intelligible framework that potentially allows for harmonization of prior findings. Current literature for example reported varying degrees of integration of the contralesional hemisphere and homotopic ROIs. Future research can interpret such variabilities in the light of the employed disconnectome approach as a potential driver as presented here.

### Lesion information towards recovery

While studies have previously discussed the diminished influence of lesion volume on the overall predictive information regarding stroke patient recovery^63^, we were able to show a clear influence of lesion volume on the structural differences between disconnectomes, as captured by spectral distance (Fig. 4B). This is particularly important in studies using chronic stroke patient data with routinely smaller lesion volumes compared to acute and sub-acute patient MRI data.^64^ Our results furthermore highlight a potential source of variability and bias in the current literature as the choice of disconnectome in conjunction with temporal dynamics of stroke lesions hold the potential to bias the outcome if not taken into consideration.

### Network properties of stroke

The application of network metrics from graph theory to brain data analysis has provided several novel insights into brain function and disease mechanisms alike, including those related to network disruptions seen in stroke.^42,65–67^ In particular, it has allowed for multidimensional groupings of stroke patients that oftentimes present with otherwise heterogeneous disease manifestations. Metrics utilized to quantify changes in network due to lesion disruption vary across the current literature and have been reviewed previously.^43^ So far, however, there has been a lack of broad consensus on feasible network metrics and the corresponding changes to them. Prior studies have for example shown how stroke induced changes to the degree of integration of the brain, as captured by shortest path length, supports prediction of 6-month stroke outcome using normative disconnectomes created from acute lesion volume masks^52^ while other studies have reported an agreement in average path length across stroke patients and healthy controls.^68^ The same holds true for small worldness^69^, a measure building upon shortest path length and the overall clustering coefficient of a network. At the same time our study showed varying degrees of importance for shortest path length as well as clustering coefficient across disconnectomes (Fig. 6B). These findings indicate the importance of varying network measures, especially in the case of subsequent machine learning based analysis, to allow for covariant information across measurements. By elevating these disagreements our study highlights the importance of a comprehensive investigation of potential biases introduced by the choice of disconnectome in future network neuroscience studies of stroke disruption.

### Capturing stroke damage in tractography

A major remaining question in the field is how damage to white matter tracts manifests and is translated to diffusion MRI. All disconnectome approaches use a binary lesion mask representing hypothetically homogeneous pathological tissue and therefore the absolute impact of stroke on the underlying brain connections. However, the true extent of damage within a given voxel of pathological tissue on the underlying white matter tracts is not well known. On the contrary, prior work has shown various influences to drive the representation of pathological tissue in MRI data. This includes the potential bias of reducing pathological tissue classification to a single modality^70^, the temporal dynamic of changes in stroke DWI data^71^ as well as the potential impact of stroke lesion on tractography without relating to the manifested disease symptom.^72^ At the same time there is a well-known dynamic regarding tissue fate in acute stroke, as highlighted by the successful integration of reperfusion therapy to alter tissue fate and improve patient outcome at the acute phase.^1^ The translation to DWI based tractography however is still an ongoing research field that was able to establish a relationship between tract integrity and functional outcome^73^ but has yet to identify an accurate interpretation of the impact of pathological voxels, as defined via a lesion mask, to concrete white matter tractography estimations across diseases.^74,75^ With this question yet to be answered it remains a strong limitation also of the current study as the assumption of capturing factual tract damage using either individual or population tractograms is only a rough estimate.

### Deep learning disease trajectories

With the current advancements in the field of machine learning and the advent of powerful deep learning architectures in particular there is a growing number of studies employing DL to the case of lesion studies, most prominently stroke. The majority of studies focused on the task of automating lesion mask estimation from various MRI modalities via image segmentation to provide time and cost-efficient clinical support.^76–79^ Several recent studies investigated the potential of deep learning models to infer stroke-specific patterns that predict recovery. These works addressed for example the problem of predicting clinical scores from deep multidimensional embeddings^18^ as well as utilizing deep architectures for specific singular tasks such as tissue outcome prediction^70^ or high dimensional lesion deficit mapping.^80^ Matsulevits and colleagues introduced the first framework to estimate disconnection networks using deep artificial neural networks^19^ which our works extends upon by introducing increased patient specificity of individual over normative disconnectomes. A graph convolutional architecture comparable to the one deployed in the present study has previously been utilized only in the case of functional MRI data for the task of lesion segmentation.^26^ Given the ability of GCN to integrate heterogeneous data in graph structures, it allowed us to create a hybrid model and integrate both normative and individual information into an informative framework encoding more than either dataset separately. Our approach represents a first step in extending our understanding of how disconnectomes can encode varying levels of information about a patient’s long term recovery potential and drive future research into precision medicine for acute stroke patients. All deep learning frameworks however require large amounts of high-quality training data and labels. This requirement presents the major limitation of the current study manifesting in two distinct challenges arising from the current data cohorts. The labels used in this study are solely based on BI. Additional measures were acquired but presented a significant number of missing values leading to a potential reduction in sample size. Additionally, the ceiling effect of the base metric BI presents a potential bias in underestimating ongoing recovery trajectory of patients. Secondly, the overall number of subjects, while presenting a comprehensive stroke population across two clinical centres, remains low for the data requirements of complex deep learning architectures. To address this, the present architecture was kept purposefully low dimensional, amassing in total only 1091180 trainable weights, and leave-one-out cross validation was applied throughout all modelling steps to retain the largest possible training set without data leakage.

## Conclusion

Our findings spotlight two major advancements towards progression of the field of MRI based stroke research and the integration of deep learning frameworks in the clinical routine. We were able to show, for the first time, far-reaching differences between modelling approaches utilizing population-derived normative and individual patient data to estimate stroke induced disconnection patterns. Future studies utilizing such disconnectomes for inference on stroke can build upon the present work by incorporating validation mechanisms to mitigate the potential impact of individual versus normative data. Our study furthermore provides a proof-of-concept for the integration of AI models into clinical diagnostics and decision making. The introduced model provides high accuracy regarding the prediction of patient specific disease trajectories, achieving performance acceptable for clinical use, while at the same time only requiring MRI data routinely available within clinical MRI stroke protocols.

## Data availability

The source code of this study is publicly available under github.com/patrikbey/SPRING-AI. The data used in this study is not publicly available due to the sensitive nature of patient MRI data. Sharing of the data is subject to the general data protection regulation (GDPR) of the European Union. For inquiries about GDPR compliant data sharing of the Charité data set the data controller Dr. Alexander Nave, Charité Berlin, Germany can be contacted. For GDPR compliant access to the UKE Hamburg data set please see the corresponding EBRAINS knowledge graph entry.^81^

## Abbreviations

AUC: area under curve
BI: Barthel index
MRI: magnetic resonance imaging
ADL: activities of daily life,
DWI: Diffusion weighted imaging
SVM: Support vector machine
DL: Deep learning,

