## supplementary for "Group-derived and individual disconnection in stroke: recovery prediction and deep graph learning"

**– Supplementary material –**

**Patrik Bey^1,2,3^**, Kiret Dhindsa^2,3^, Torsten Rackoll^4,5,6^, Jan Feldheim^7^, Marlene Bönstrup^7^, Götz Thomalla^7^, Robert Schulz^7^, Bastian Cheng^7^, Christian Gerloff^7^, Matthias Endres^2,3,6,8,9,10^, Alexander Nave^4,8,9,10^, and Petra Ritter^2,3,11,12,13^

Author affiliations:
1 High Dimensional Neurology Group, UCL Queen Square Institute of Neurology, University College London, Russell Square House, Bloomsbury, London, UK
2 Berlin Institute of Health at Charité, Charitéplatz 1, 10117 Berlin, Berlin, Germany
3 Department of Neurology with Experimental Neurology, Brain Simulation Section, Charité – Universitätsmedizin Berlin, corporate member of Freie Universität Berlin and Humboldt-Universität zu Berlin, Berlin, Germany
4 Center for Stroke Research Berlin, Charité – Universitätsmedizin Berlin, Berlin, Germany
5 Berlin Institute of Health QUEST Center for Responsible Research Berlin, Charité – Universitätsmedizin Berlin, Berlin, Germany,
6 Klinik und Poliklinik für Neurologie, Kopf und Neurozentrum, Universitätsklinikum Hamburg-Eppendorf, Hamburg, Germany
7 Klinik Und Hochschulambulanz für Neurologie, Charité – Universitätsmedizin Berlin, Berlin, Germany,
8 German Centre for Cardiovascular Research, Partner Site Berlin,Berlin, Germany,
9 German Center for Neurodegenerative Diseases, Partner Site Berlin, Berlin, Germany,
10 German Center for Mental Health, Partner Site Berlin, Berlin, Germany
11 Bernstein Focus State Dependencies of Learning and Bernstein Center for Computational Neuroscience, Berlin, Germany
12 Einstein Center for Neuroscience Berlin, Berlin, Germany
13 Einstein Center Digital Future, Berlin, Germany

### Data requirements

To enable disconnectome creation for each patient the underlying MRI data was processed (see paragraph MRI preprocessing below) resulting in the requirement of certain modalities as given in Supplementary Table 1. MRI data was included for patients from acute to early sub-acute state. For each patient anatomical scans including MPRAGE T1 weighted scans as well as Fluid Attenuated Inverse Recovery (FLAIR) scans were acquired. Based on the structural modalities stroke lesion masks were manually drawn. The lesion mask filename contains information about the underlying modality used for segmentation (i.e. {Mod} = T1w). In addition, single phase encoding diffusion weighted imaging (DWI) data was acquired for each patient individually with corresponding b-vector and b-value files.

**Supplementary Table 1 Data requirements** to perform the presented processing and downstream analysis presented in this study.

| **BIDS filename** | **notes** |
| --- | --- |
| sub-{ID}_T1w.nii.gz | Anatomical full brain MPRAGE image |
| sub-{ID}_FLAIR.nii.gz | Anatomical full brain FLAIR image |
| sub-{ID}_{Mod}_lesion_mask.nii.gz | Binary lesion mask in {Mod} space, e.g. FLAIR |
| sub-{ID}_dwi.nii.gz | Diffusion image volume |
| sub-{ID}_bval.bval / _bvec.bvec | Corresponding b-value and orientation files |

Such a comprehensive data set for each patient enabled the downstream processing as described below. A description of MRI acquisition parameters can be found in prior publications describing the respective data sets.^1,2^

Additionally, this study required clinical assessment of the patient’s disability at initial sub-acute timepoint and three months post stroke incident via Barthel Index.

### Processing framework

The MRI data utilized in this study was pre-processed to enable downstream analysis including the creation of both individual and normative disconnectome. The raw MRI data was standardized according to BIDS^3^ to allow utilization of the existing stroke specific processing pipeline LeAPP.^4^ In a first step structural MRI data was pre-processed using LeAPP’s PreFreeSurfer step including stroke specific processing steps such as cost function masking^5^ and enantiomorphic normalization.^6^ In a second step the derivatives were run through the FastSurfer^7^ pipeline to enable fast and accurate surface reconstruction and segmentation. Subsequent structural processing included the PostFreeSurfer and parcellation mapping step of LeAPP resulting in individual brain parcellations for each patient. Diffusion processing was then performed utilizing LeAPP’s DWI processing pipeline resulting in individual tractograms for each patient containing a total of 10 million tracts. Such derivatives enabled the creation of disconnectomes as described in the main manuscript of this study.

### Network measures

The following measures were included in the current analysis and are briefly described here. For a more in-depth analysis of the metrics please see e.g. the works by Rubinov and Sporns^8^, Newmann^9^, and Bassett and Sporns^10^.

**Supplementary Table 2 Network metrics** describing both disconnectome graphs as utilized in the study.

| **Network metric** | **Information captured** |
| --- | --- |
| *Node strength* | Weighted variant of the node degree, the basic measure for node importance, incorporating the strength of connected edges. |
| *Clustering coefficient* | Describing the degree of local clustering across the whole network. |
| *centrality* | Average betweenness centrality, capturing the importance of a given node in connecting distant nodes of the network. |
| *assortativity* | Capturing resilience of a network as defined by the mutual connectivity of nodes across all links. |
| *density* | A single measure describing the relationship of present connections over all possible connections for a given network. |
| *Shortest path length* | Encoding the average shortest path across all connections as a measure of integration of information exchange between nodes. |
| *Non-randomness* | A measure describing the deviation of the given graph to random Erdös-Rényi graphs with the same set of nodes. |
| *transitivity* | Global value of clustering without bias on local nodes with a low degree. |
| *diameter* | The longest path between any connected node pair of a network |
| *periphery* | Count of number of nodes with edges whose length is equal to the diameter of the network providing a measure for the integration of distant nodes into the network. |
| *Bridge count* | Providing the number of edges in a graph whose removal would increase the number of connected components in our case creating sub-graphs of disconnected ROIs. |
| *Independence count* | A measure to validate the number of ROIs in the network without connections to each other in the initial graph, providing information about the spread of the graph across all potential node of the brain parcellation. |
